# Intra-arterial transplantation of autologous mesoangioblasts in m.3243A>G mutation carriers is safe – first phase I/II human clinical study

**DOI:** 10.1101/2025.01.17.24319434

**Authors:** Florence H.J. van Tienen, Janneke G.J. Hoeijmakers, Christiaan van der Leij, Erika Timmer, Nikki Wanders, Patrick J. Lindsey, Fangzheng Yi, Fong Lin, Susanne P.M. Kortekaas, Helene Roelofs, Inge M. Westra, Pauline Meij, Lambert A.C.M. Wijnen, Irenaeus F.M. de Coo, Hubert J.M. Smeets

## Abstract

Progressive myopathy and exercise intolerance significantly impair quality of life in over 50% of m.3243A>G mutation carriers, with no curative therapy currently available. We hypothesize that intra-arterial administration of autologous, mtDNA mutation-free myogenic stem cells, mesoangioblasts, can reduce mutation load, enhance oxidative phosphorylation, and improve muscle function. To test this, the tibialis anterior muscles of three m.3243A>G mutation carriers were damaged by eccentric exercise before infusion of 50 million /kg autologous mesoangioblasts into the left anterior tibial artery. The right tibialis anterior muscle served as control. Expanded mesoangioblasts had a mutation load of <15%, though culturing increased this by 7–15%. Infusion caused mild, transient discomfort without serious adverse events or vascular obstructions, as confirmed by angiography. Blood and muscle biopsies revealed no systemic or local inflammation at 24 hours and 4 weeks post-transplantation. Biopsies of the treated muscle suggested mesoangioblast migration and early signs of regeneration. This first-in-human study demonstrates that intra-arterial administration of autologous mesoangioblasts is safe, with promising, though inconclusive, evidence for muscle regeneration and mesoangioblast homing. These findings support further investigation into the therapeutic potential of mesoangioblasts for treating myopathy in m.3243A>G mutation carriers.

## Introduction

Mitochondrial myopathies (MM) represent a spectrum of progressive muscle diseases caused by compromised energy production in the mitochondrial oxidative phosphorylation (OXPHOS) system [1]. In 20-25% of the cases, MM are caused by a heteroplasmic mutation in the mitochondrial DNA (mtDNA) [2], which is characterized by co-existence of both wild-type and mutated mtDNA copies. One of the most frequent heteroplasmic mtDNA mutations is the m.3243A>G mutation in the dihydrouridine loop of mitochondrial tRNA Leu(UUR) gene [3]. Skeletal muscles are commonly affected in symptomatic m.3243A>G mutation carriers. More than 90% of the m.3243A>G patients display ragged-red fibers, and lactic acidosis in muscle and progressive myopathy and exercise intolerance occur in more than half of the m.3243A>G patients with a mean heteroplasmy level of ∼50% (range 30%-74%) in skeletal muscle [4–6]. Age-related muscle loss even worsens the clinical manifestations, since the percentage of individuals with myopathy and severity of myopathy among m.3243A>G patients drastically increases with age and has highest incidence in the fifth decade of life [5]. Severe fatigue complaints were also reported by the majority (77%) of clinically affected m.3243A>G mutation carriers [7]. Patients report muscle fatigue and weakness as having the greatest impact on quality of life [8, 9]. Consequently, treating the muscle-related complaints, irrespective of other possible disease manifestations, would be highly beneficial for patients.

Myogenic stem cell-based therapies are highly promising to combat myopathy and exercise intolerance. Myogenic stem cells called mesoangioblasts (MABs), are currently the only myogenic precursors that fulfill all criteria to be used as advanced therapy medicinal product (ATMP) for systemic treatment, namely good *ex vivo* proliferation capacity, high myogenic capacity and a capability to cross blood vessels, allowing intra-arterially (systemic) delivery towards affected muscle. Successful preclinical studies showed that treatment with three doses of 50 million/kg ex-vivo expanded MABs resulted in significant numbers of dystrophin positive muscle fibers and clinical improvement in mice and dog models of Duchenne Muscular Dystrophy (DMD) [10–12].

Subsequent, intra-arterial delivery of allogeneic MABs in DMD boys in a first human phase I/II clinical study demonstrated that the treatment was relatively safe, and that dystrophin positive fibers were detected, but not sufficient for functional improvement. As MABs are not immune privileged, usage of immunosuppressive agents was required upon transplantation of allogeneic MABs, which might have a negative effect on homing of the MABs [13]. The use of autologous MABs eliminates the use of immunosuppressive agents, but requires availability of MABs with no or a low mutation load. We previously demonstrated for heteroplasmic m.3243A>G mutation that six out of nine MABs cultures displayed a mutation load below 15% [14], enabling direct *ex vivo* expansion of patient derived healthy MABs for autologous cell therapy to combat MM.

Here we present our first in human phase I/II clinical trial of autologous MABs as ATMP. The primary goal was to assess safety of one administration of 50 million/kg autologous MABs in the tibial artery of three patients heteroplasmic for the m.3243A>G mutation (NCT05063721 clinicaltrials.gov). Since carriers of mtDNA mutations typically do not experience muscle wasting and associated regeneration, as seen in DMD, and MABs require muscle damage and inflammation for extravasation [10, 15], a bout of eccentric exercise was performed 24 hours prior to transplantation. This eccentric exercise induces muscle damage, infiltration of leukocytes and expression of inflammation markers [16, 17]. Following administration of the MABs, participants were observed in the hospital for 24 hours post-infusion and (serious) adverse events (S(AEs)) were documented. Pre- and post-infusion angiography were performed. Plasma CK, CK-MB and LDH were analyzed as markers of (skeletal muscle) tissue damage. Plasma and tibialis anterior muscle inflammation was assessed using the pleiotropic cytokine IL-6, which is a key mediator in acute-phase response to infections and injuries, the early response cytokine TNFα that is mostly involved in localized tissue inflammation, and SDF1a as it is a highly effective chemoattractant, which activate leukocytes and is induced by pro-inflammatory cytokines, such as TNFα.

Our secondary goal was to assess preliminary effectiveness with respect to skeletal muscle homing and regeneration. MABs can be discriminated in a muscle biopsy by the expression of pericyte markers, such as Alkaline phosphatase (AP) and Neural glia antigen-2 (NG2) [12]. However, upon autologous transplantation, the transplanted cells cannot be discriminated from the residing MABs. Indocyanine green (ICG) dye is an FDA-approved dye that is widely used in vivo with near infrared light [18]. In this study, ICG labeling of MABs prior to infusion was tested to enable migration assessment of infused MABs in tibialis anterior muscle biopsies of treated and untreated legs 24-hours after administration. For reasons explained in the results part, this was done in only 1 patient. In addition, NCAM staining was used to quantify formation of new muscle fibers within these muscles 4-6 weeks after ATMP administration.

## Materials and Methods

### Study design

This intra-subject controlled mono-center study, approved by the Central Committee on Research Involving Human Subjects (CCMO) in the Netherlands (NL68732.000.19, June 7, 2020), was conducted according to the principles of the Declaration of Helsinki and in accordance with the Medical Research Involving Human Subjects Act (WMO) and other guidelines, regulations and acts. Carriers of the m.3243A>G mutation were enrolled in this phase I/II clinical study at Maastricht UMC (MUMC) in the Netherlands and written informed consent was obtained before execution of the study. This study comprised five study visits (see figure 1 for overview study design). Visit 1 was a screening visit to assess eligibility for participation. At this visit, in- and exclusion criteria, virology (HIV/Hepatitis/HTLV/TPHA) and medical history were evaluated, a neurological and physical examination was performed, and a skeletal muscle biopsy was obtained from the vastus lateralis muscle to determine if the m.3243A>G mutation load in MABs was below 15%. At visit 2, a skeletal muscle biopsy from the vastus lateralis muscle was collected from eligible participants for isolation and expansion of autologous mesoangioblasts at the GMP facility of the Leiden University Medical Center (LUMC). Upon completion of MABs expansion, visit 3 and 4 were scheduled. At visit 3, one day before transplantation, a maximal bout of eccentric exercise was performed with both lower legs to induce muscle damage and inflammation. The next day (visit 4), autologous mesoangioblasts were administered into the tibialis anterior of the left leg via a catheter in the femoral artery. Pre- and post-administration angiography was performed, and the participants were observed for 24-hours, vital signs were recorded (bi)hourly, and venous blood samples were collected every 8 hours. Twenty-four hours after transplantation, a tibialis anterior muscle biopsy of both the left (treated) and right (untreated) leg was collected. Patients returned to the hospital four to six weeks later for the last study visit (visit 5). At this study visit, a physical and neurological examination was performed, a venous blood sample was collected and a tibialis anterior muscle biopsy of both the left (treated) and right (untreated) leg was collected. The following paragraphs describe all procedures used.

**Figure 1.**
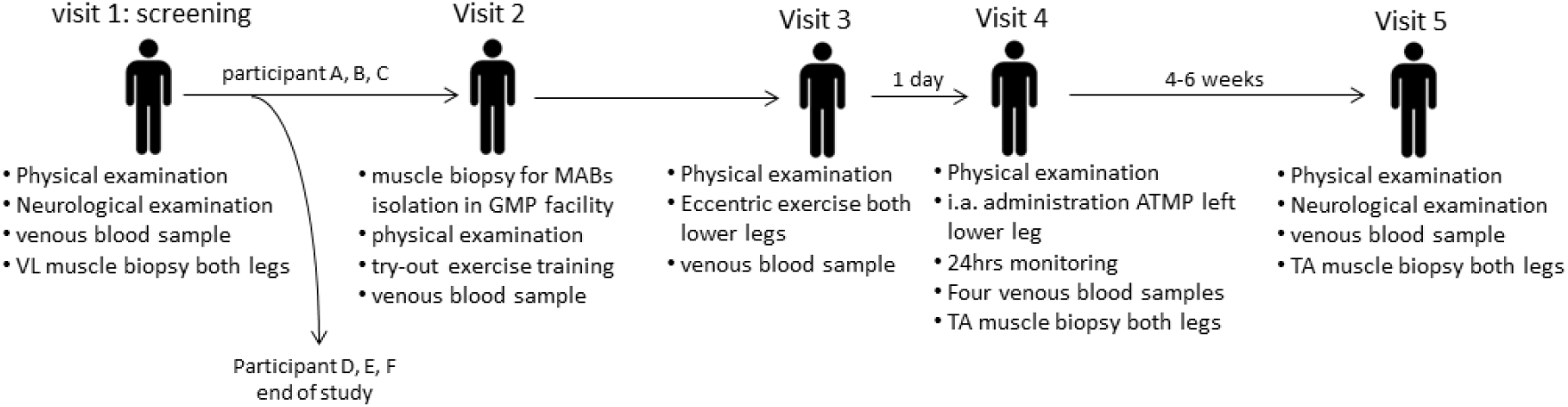
Study design. Study design including interventions at each study visit. TA = tibialis anterior; VL = vastus lateralis.

### Clinical and neurological examination

A baseline routine clinical examination was performed at each study visit to record the body height and weight, the blood pressure, and the heart rate at rest. A neurological clinical examination was performed at the first and last visit to document the disease course and included interval medical history, present illness and general medical history; Eye Motor and Verbal response score (EMV); global mental status; cranial nerves; muscle strength (MRC sum score) muscle tone and bulk; gait testing; sensory and coordination examination; reflex testing.

### Skeletal muscle biopsies

The vastus lateralis and tibialis anterior skeletal muscle biopsies were collected using the 14G Pro-Mag I automatic biopsy instrument. First, the skin was locally anesthetized using 2-3 ml lidocaine and a 0,5 cm incision was made in skin and fascia after which the muscle samples were collected using the Pro-Mag I automatic biopsy instrument. Following the muscle biopsy, the skin was closed using a Steristrip® and covered by a Tegaderm®. The vastus lateralis muscle samples (∼100mg) collected for mesoangioblast culture were transferred sterile into a 50ml tube containing 20 ml IMDM medium supplemented with 50µg/ml gentamycin, 1% insulin-transferrin-selenium, 1% non-essential amino acids, 0.1 mM 2-mercaptoethanol and 10ng/ml FGF. The tibialis anterior muscle biopsies were embedded in Tissue-Tek® and subsequently snap frozen in liquid nitrogen cooled isopentane.

### Mesoangioblast isolation, expansion, ICG labelling and medicinal product formulation for clinical administration

The sterile collected muscle tissue, the starting material of the production process, was transported cooled (2-8°C) to the GMP facility. Following reading of the temperature logger, checking of the transport documents and initiation of sterility tests, the start material was dissected into small pieces (±2 mm) using a surgical knife and fat was removed when present. Selected fragments were transferred into type I human collagen (Symatese) coated 10cm dishes containing 4 ml complete MABs medium (IMDM medium supplemented with 10% FBS, 50µg/ml gentamycin, 1% insulin-transferrin-selenium, 1% non-essential amino acids, 0.1 mM 2-mercaptoethanol and 10ng/ml FGF) and were incubated at 37°C/4%O2/5%CO2. After ∼1-week, preliminary growth of adherent cells will become visible. A characteristic of pericytes is that they are initially apparent as small, round, very refractile cells that are floating or weakly adhering to the layer of flat cells below. After ∼2 weeks, the culture medium and floating fragments (cells/pieces of muscle tissue) were carefully discarded, the adherent cells were detached using Tryple select and reseeded at 10,000-12,000 cells/cm2 in MABs medium in a new flask. One day and two days later, the floating cells (=pericyte-derived progeny: MABs) were recovered and transferred to new flasks at density 4,000-6,000 cells/cm2. MABs were expanded, CD56+ cells were depleted using CliniMACS CD56 reagent (Miltenyi Biotec) and the intermediate product (IP) was frozen in 90%FBS/10%DSMO at a concentration of 1×10E6/ml. IP testing comprised immunophenotyping (CD13+, CD44+, CD31-, CD34-, CD45- and CD56-) using a FACS Canto II (BD). Viability and cell number quantification was obtained using Trypan blue staining and hemacytometer counting. m.3243A>G mutation load analysis was performed as described below. Patient identity verification was checked using AmpFLSTR kit and LAL assay for endotoxin. Mycoplasma PCR test and Bactec test for microbiological control of cellular product were performed by the department of Clinical microbiology LUMC. One to two weeks before infusion, IP was thawed, expanded for 1-2 passages and medicinal product (MP) was formulated one day before infusion. The MP consisted of either unlabeled, or 90% unlabeled and 10% labelled with IC-Green (Verdye) at a concentration of 30ug/ml in culture medium for minimum 1 hour and maximum 3 hours before harvesting. After harvesting, the autologous MABs were washed with NaCl with albumin and the required number of MABs (50 million x 5% of body weight for treatment of one lower leg) was re-suspended in Hypothermosol (BioLife solutions) and 5IU/ml heparin (5×10E6/ml). The MP was stored and transported to MUMC between 2-8°C till infusion.

### Quantification of m.3243A>G mutation load

For quantification of the m.3243A>G mutation load in mesoangioblasts and skeletal muscle, DNA was isolated using the Promega wizard DNA extraction kit according to manufacturer’s instructions. Quantification of the m.3243A>G mutation load was performed using Genescan fragment analysis of *HaeIII* digested PCR products, which results in a 48bp fragment in case of m.3243A>G mutation and a 122bp fragment for wild-type mtDNA, as described [14]. Alternatively, next generation sequencing of m.3243 containing PCR product was performed using a PacBio analyser at the department of Clinical Genetics of Maastricht UMC.

### Eccentric exercise of tibialis anterior muscles

A bout of eccentric exercise training of both lower legs was performed on a Biodex isokinetic dynamometer to induce muscle damage and inflammation of the m. tibialis anterior. During exercise training, patients positioned in supine lying position, where resistance was applied to the dorsum of the foot just above the toes to resist dorsiflexion, while stabilization was applied to the lower leg. The amount of resistance increased gradually to be suitable to the participant tolerance. Each subject rested for thirty seconds after each set of 10 contractions to avoid fatigue. Participants performed as many sets of 10 contractions as feasible for them.

### Intra-arterial administration of MP, angiography and 24h monitoring

The MP was intra-arterially delivered to the anterior tibial artery (ATA) of the left lower leg via a catheter in the left femoral artery. Following local anesthesia with 10 ml lidocaine 1%, a 4 Fr introducer sheath was inserted antegradely in the common femoral artery followed by injection of 3.000 units of heparin through the sheath. Initial digital subtraction angiography (DSA) targeted on the vessels of the lower leg (6 ml of 50% diluted contrast agent with 3 ml per second injection rate) was performed. The ATA was identified and selectively catheterized with a 2.7 Fr micro-catheter (Progreat Terumo) using a micro-guidewire (0.021”). Then the ATMP was injected at a rate of 1-2 ml/min in the target vessel under continuous fluoroscopy. After finishing the procedure, post-injection DSA was acquired to assess patency of the culprit vessels. The sheath was removed, and the puncture location was manually compressed for at least 10 minutes. Subsequently, the patient was immobilized for at least 4 hours and monitored for 24 hours in the hospital after infusion. Each hour, the participants temperature, blood pressure, heart rate, oxygen saturation and breathing frequency per minute were recorded, as well as pain score (0 being no pain and 10 being very much pain), EMV score (maximal score 4-6-5), MRC strength score of both legs (0-5) and visual inspection of the legs.

### Venous blood sampling and analysis

At study visits 1, 2, 3, and 5, one venous blood samples was collected and four samples, every eight hours, were collected during 24h monitoring at visit 4. Upon collection of venous blood, samples were centrifuged (20 minutes at 2000 RPM) and plasma was either analyzed directly or stored in cryovials at −80°C till analysis. Analysis of Creatinine, Creatine Kinase (CK), CK-MB, and lactate were performed by the central diagnostics laboratory (CDL) of Maastricht UMC. Analysis of Hepatitis B/C, HIV, HTLV, and TPHA were performed by the department of microbiology and virology of Maastricht UMC. Plasma levels of TNFα, SDF-1, and IL-6 were analyzed using ELISA according to manufacturer’s protocol (R&D systems) and signal intensity was assessed using a CLARIOstar microplate reader (BMG Labtech).

### (Immuno)stainings of skeletal muscle biopsies

Skeletal muscle biopsies of m. tibialis anterior were upon collection snap frozen in OCT (Tissuetek) in isopentane cooled using liquid nitrogen, and stored at −80°C. Muscle cross-sections of 7 μm were made using a cryotome (Leica) and directly frozen at −20°C. Immunostainings, cryosections were fixated with 4% paraformaldehyde (Sigma-Aldrich) for 10 minutes, followed by 3 times wash with PBS. To assess homing in biopsies collected 24 hours after administration, cryosections were incubated for 90 minutes in blocking solution (PBS containing 2% bovine serum albumin, 5% fetal bovine serum, 0,2% Triton X-100, 0,1% sodium azide), three times washed with PBS and incubated overnight with primary antibody anti-NG2 (MAB5384-1 Millipore Sigma) in a blocking solution at 4°C. After overnight incubation, the cryosection was washed three times with PBS and incubated with 1:500 Alexa Fluor 594 goat-anti-mouse secondary antibody and 1:500 WGA 488 goat-anti rabbit (ab73593 Abcam) for one hour at room temperature. Nuclei were stained with DAPI in glycerol. Slides were imaged on a Olympus BX51 fluorescent microscope. Images of NG2 and WGA staining were merged using Image J and the number of NG2+ cells and number of muscle fibers were counted manually. For quantification, slides were randomized and counting was performed by a blinded researcher. A minimum of six fields of view were quantified per sample. To assess formation of new muscle fibers in muscle biopsies collected 4-6 weeks after administration, cryosections were incubated for 60 minutes in blocking solution (PBS containing 2% bovine serum albumin, 5% fetal bovine serum, 0,2% Triton X-100, 0,1% sodium azide), three times washed with PBS and incubated overnight with 1:50 primary antibody anti-NCAM (5.1H11 DSHB) in a blocking solution at 4°C. After overnight incubation, the cryosection was washed three times with PBS and incubated with 1:100 rabbit-anti-mouse PO secondary antibody (Agilent DAKO) in a blocking buffer for one hour at room temperature. After washing three times with PBS, samples were incubated with 1:50 DAB Chromogen (Agilent DAKO, K3467) in a substrate buffer for 10 minutes at room temperature. After a wash with demiwater, nuclei were stained using hematoxilin for two minutes, washed with water for five minutes, dehydrated and covered with a coverslip with Entellan. Samples were imaged using a 3D Histech Panoramic 1000 slide scanner. The number of NCAM+ cells and total number of muscle fibers were counted manually using 3D Histech Caseviewer software v2.4. For quantification, slides were randomized and counting was performed by a blinded researcher.

### Quantitative PCR

Total RNA was isolated from the tibialis anterior muscle biopsies using TRIzol reagent, and purified with the High pure RNA isolation kit (Roche). cDNA was synthesized from 1ug RNA using qSCRIPT reagent according to manufacturer’s protocol (QuantaBio). Quantitative PCR was performed on a Roche LC480 in 10ul reactions containing 1x Sensimix SYBR Hi-Rox (Bioline), 1.5uM forward and reverse primer (see Supplementary Table S3 for primer sequences) (IDT) and 5 ng cDNA, using the following cycling conditions: an initial step of 10 minutes at 95°C, followed by 40 cycles of 15 seconds at 95°C and 1 minute at 60°C. The mRNA levels of each gene were normalized to those of the housekeeping gene TATA-box binding protein (TBP).

### Statistics

A paired Student t-test was used to assess if the number of NG2+ positive cells and number of NCAM+ muscle fibers was significantly different between m. tibialis anterior of the treated leg compared with the untreated leg (p<0.05 was considered significantly changed). For all other data, descriptive statistics was used.

## Results

### Study population

From November 2020 to May 2022, six m.3243A>G mutation carriers were enrolled in this study and completed the screening visit (visit 1), which consisted of a clinical and neurological examination, checking in- and exclusion criteria, describing medical history, and obtaining a skeletal muscle biopsy from the m. vastus lateralis for analysis of the m.3243A>G mutation load in MABs. Five out of six participants were eligible for participation based on their m.3243A>G mutation load in MABs being <15% (Table 1). Patient D displayed a m.3243A>G mutation load of 19% in MABs and therefore her study participation ended after visit 1. Also, participant E was excluded from further participation because of a vascular obstruction in his left leg, and participant F was excluded based on investigator’s opinion with respect to his ability to understand and follow study procedures. Participants A, B, C were included and completed the clinical study (visit 2 – 5).

**Table 1.** Subjects included in phase I/II clinical study.

| Id | Age<br>(years) | Gender | Weight<br>(kg) | m.3243A>G %<br>sk. muscle* | m.3243A>G<br>% MABs<sup>#</sup> | MRC sum<br>score | Clinical<br>phenotype | Status |
| --- | --- | --- | --- | --- | --- | --- | --- | --- |
| **A** | 50-55 | F | 65-70 | 63,1 | 6,7 | 110 | Diabetes Mellitus | Completed all 5<br>study visits |
| **B** | 30-35 | F | 55-60 | 55,0 | 7,5 | 110 | Myalgia after<br>exercise | Completed all 5<br>study visits |
| **C** | 50-55 | M | 65-70 | 58,8 | 9,5 | 106 | Diabetes<br>Mellitus,<br>hypertension,<br>small fiber<br>neuropathy.<br>Weakness hip<br>flexors and knee<br>flexors MRC<br>score 4. | Completed all 5<br>study visits |
| **D** | 40-45 | F | 55-60 | 68,0 | 19 | 110 | Hypertension.<br>Exercise<br>intolerance. | Excluded after<br>visit 1 based on<br>mutation load in<br>MABs >15% |
| **E** | 65-70 | M | 70-75 | 65,0 | 6,5 | 110 | Stenosis left common iliac artery and superficial femoral artery | Excluded after visit 1 due to vascular obstruction left leg |
| **F** | 35-40 | M | 50-55 | 57,0 | 8,6 | 110 | Diabetes Mellitus | Excluded after visit 1 based on opinion investigators |

\* vastus lateralis skeletal muscle biopsy; # analyzed at visit 1; F=female, M=male; MRC sum score = Medical Research Council sum score, maximum score is 110.

### Autologous mesoangioblast medicinal cell products

In the GMP facility of the LUMC, mesoangioblasts were isolated and expanded from vastus lateralis muscle biopsies taken from subjects A, B, and C for the preparation of the medicinal product for infusion. After isolation and expansion, a batch of MABs was frozen and analysis of the intermediate product release criteria was performed. All intermediate product criteria were met, only the MABs m.3243A>G mutation load of subject C was out of specification, namely 16%, while criterion was ≤15%. With consent of participant C, intermediate product was used for generation of the MP for clinical administration. One week before the scheduled date of intra-arterial transplantation, the MABs intermediate product was thawed and expanded. In order to enable detection of the transplanted MABs in the muscle biopsy collected 24 hours after ATMP transplantation, 10% of the autologous MABs of participant B were labelled with IC-green (ICG) before harvesting. In this case, the ATMP consisted of 148 million (90%) unlabeled MABs and 14,8 million (10%) ICG-labeled MABs in 29 ml Hypothermosol (5 million/ml) and 5U/ml Heparin. As explained in the results section, for participant A and C, the medicinal product consisted respectively of 175 and 168 million unlabeled autologous MABs in 35 and 34 ml Hypothermosol (5 million/ml) and 5U/ml Heparin. All three MPs fulfilled conditional release criteria (Table S1a), were QP released at the GMP facility, then transported to MUMC and stored both at 2-8°C until administration. All three administrations took place within the allotted 30-hour window after formulation. Final MP release criteria that only became available after administration consisted of mycoplasma, final microbiological control of cellular product, endotoxin, patient verification and m.3243A>G mutation load analysis (Table S1b). Except for m.3243A>G mutation load analysis, all were within specification. The m.3243A>G mutation load criterion was set at ≤15%, but were respectively 18%, 27% and 23% for subjects A, B and C. Participants A and C showed on average a 7-8% increase between IP and mean MP m.3243A>G mutation load analysis. The increase in participant B of the clinical study was 15%, resulting in a mutation load of 27% in the MP. In addition, large variation (mean difference 6%) was observed upon analysis of MP mutation load using two different methods, namely NGS Pacbio analysis and Genescan fragment analysis (Supplementary table S4).

### Primary objective: Safety of intra-arterial administration of autologous MABs

As this was the first in human clinical administration of autologous mesoangioblasts, safety assessment was the primary study objective. Pre- and post-administration angiography verified that there were no vascular obstructions in the left lower leg following autologous mesoangioblast administration. In none of the three participants, drastic changes or values outside the normal range were observed of blood pressure, heart rate, breathing frequency and oxygen saturation during 24 hours monitoring following transplantation (Table 2 and Figure 2). From all participants, an ECG before and 24h after ATMP administration demonstrated no changes for the heart.

**Figure 2.**
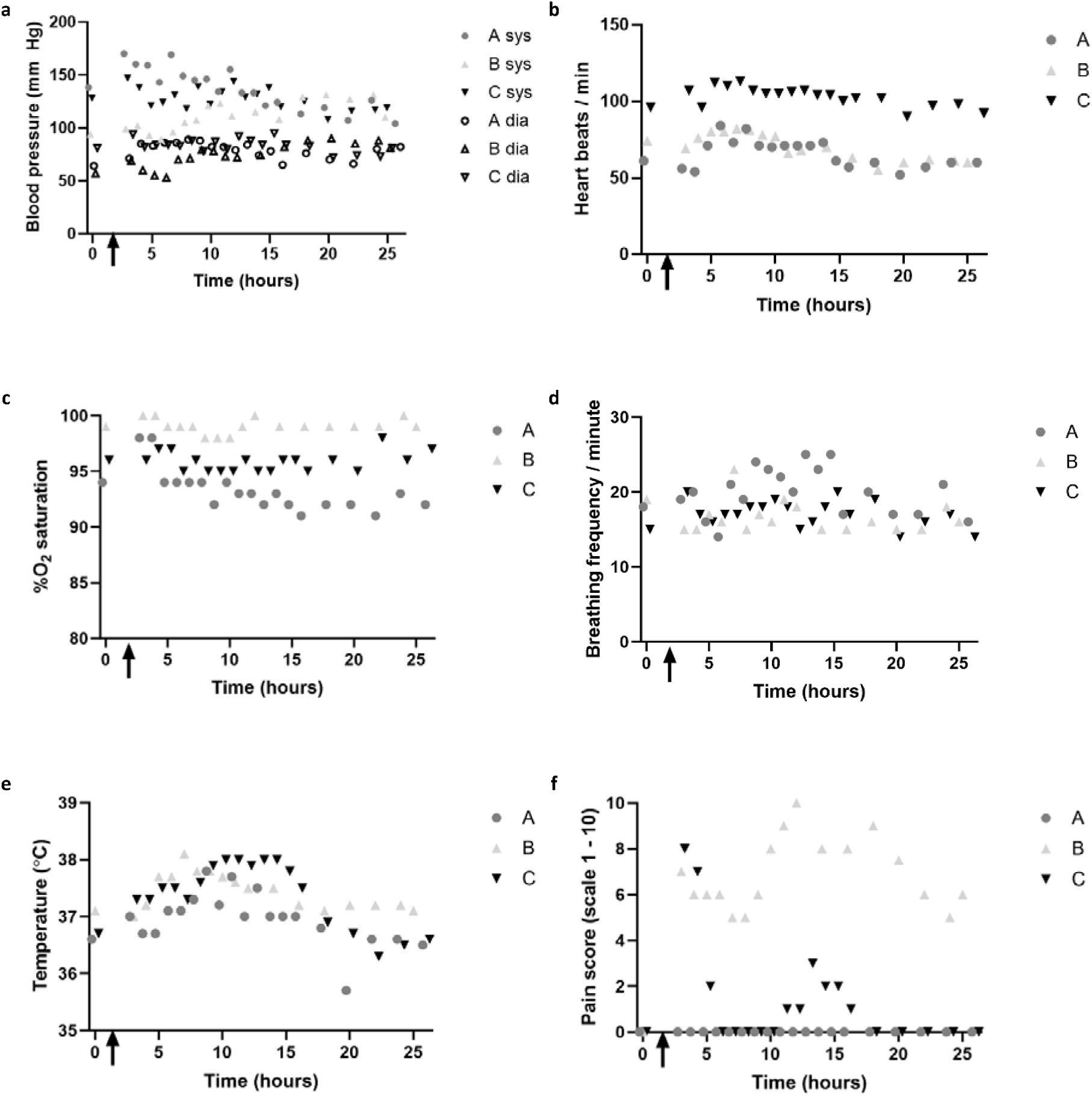
Monitoring of vital signs during 24h after ATMP delivery After base-line measurements at t=0, ATMP was delivered at t=1-2h, and hourly monitoring of blood pressure during 24hr observation at visit 4 of (a) Blood pressure; Heart rate (b); %O_2_ saturation (c) and breathing frequency (d), temperature (e) and pain score (f) were assessed before (t=0) and every hour during 24 hour observation following ATMP delivery; The arrow indicates ATMP delivery.

**Table 2.** Summary of (S)AEs and vital signs during 24h monitoring following MABs administration.

| ID | SAE | AE | BP (mm HG) |  | HR/min | %O<sub>2</sub> | BF/min |
| --- | --- | --- | --- | --- | --- | --- | --- |
|  |  |  | SYS | DIA |  |  |  |
| **A** | No | No | 137±20 | 79±8 | 66±9 | 93±2 | 20±3 |
| **B** | No | Grade II, rash, and redness TA muscle left leg | 111±14 | 74±9 | 70±9 | 99±1 | 17±2 |
| **C** | No | No | 128±10 | 83±6 | 103±6 | 96±1 | 17±2 |

SAE: serious adverse event; AE: adverse event; BP: blood pressure, SYS: systolic BP; DIA: diastolic BP; HR/MIN: heart rate/minute; %O<sub>2</sub>: percentage O<sub>2</sub> saturation; BF/MIN: breathing frequency per minute; Data BP, HR/MIN, %O<sub>2</sub> and BF all shown as mean±stdev during 24 hour observation.

All three subjects completed the five study visits, and no SAE or AE was observed between visit 2 and 5 in the 28 days following ATMP delivery. In subject B, a local grade II AE was observed during the ATMP administration. This local AE started as a local skin rash with itching, and redness of the skin on the front side of the left lower leg and was in hours after the procedure followed by soreness and mild swelling of the *m. tibialis anterior*. Participant B was discharged as scheduled after 24-hour monitoring, received Ibuprofen (400mg) pain medication and an extra visit was scheduled for inspection of the leg. No 24hr muscle biopsies were taken. When the participant visited the out-patient clinic three days later, the rash had largely disappeared, redness of the skin was reduced, and soreness was less. Participant B received 90% unlabeled and 10% ICG-labeled MABs as ATMP. As it was not possible to assess if the adverse event was due to the use ICG or due to a sensitivity of the patient to other components of the ATMP or the administration procedure, we decided to leave out ICG in subsequent ATMP to eliminate a variable for safety as primary endpoint, which will not be present anyhow in the eventual therapeutic procedure. In both participant A and C, intra-arterial administration of the unlabeled MABs as ATMP was generally well tolerated. Participant A experienced discomfort in the treated left lower leg during administration, which improved by moving her feet and was completely resolved after ATMP administration was completed. Participant C experienced pain in the left leg towards the end of the intra-arterial ATMP administration and during the 10-minute pressure on thigh by the intervention radiologist upon removing catheter from the femoral artery. Paracetamol was given, and pain was nearly gone within couple of hours after the procedure. The front side of the treated m. tibialis anterior was a bit warm in participant A and C, and also a bit red in participant C, during the first hours after the procedure.

To assess systemic inflammation due to ATMP delivery, multiple blood samples were collected, of which two before ATMP administration (t=-24h and t=0h), and four after administration (t=8h, 16h, 24h and 4-6 weeks). In these blood samples, IL-6, CK, CK-MB, LDH, TNFα, SDF1a were analyzed (Figure 3). IL-6 plasma level was elevated at blood sampling 8 hours after administration but returned to baseline within 24 hours after ATMP administration. This ceased within a couple of hours after administration, when administrated cells were cleared from the circulation, causing cessation of the IL-6 signaling cascade. In line with this, LDH, CK and CK-MB were not elevated in participant A and C. Only in participant B, who experienced the adverse event, CK and CK-MB were increased 24h after ATMP administration, which was back to normal at the last study visit. The same was observed for TNFα. Lastly, ATMP administration did not impact plasma SDF1a levels. Lack of SDF1a activation is in line with the other parameters, verifying that autologous MABs delivery as ATMP is not associated with a systemic inflammation or immune response.

**Figure 3.**
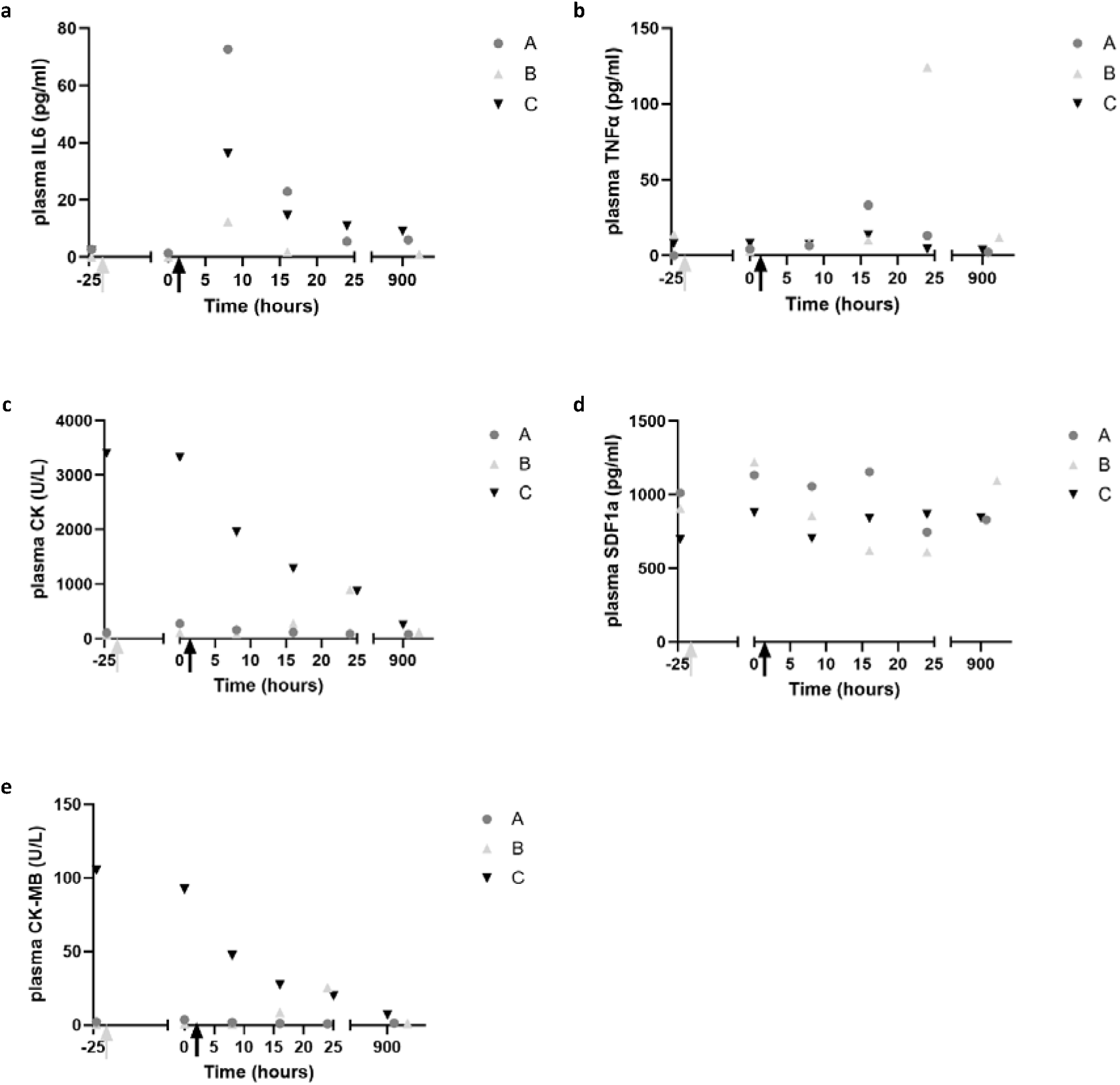
Assessment of body temperature, pain score, and markers of inflammation and tissue damage in venous blood samples following ATMP delivery. ATMP administration was performed at t=1-2h, venous blood samples were collected before ATMP administration at visit 3 (t=-24 hour) and at start of visit 4 (t=0 hour), three times after ATMP delivery at visit 4 (t=8, 16 and 24 hour), and at visit 5 (4-6 weeks after ATMP delivery and were analyzed for IL-6 (a); TNFa (b); CK (c); SDF1a (d) and CK-MB (e). Grey arrow indicates time point eccentric exercise bout lower legs at visit 3 and black arrow indicates time point ATMP infusion at visit 4.

To assess if autologous MABs administration resulted in inflammation of the tibialis anterior muscle, H&E staining and gene expression analysis of inflammation markers TNFα, IL-6, SDF-1 was performed. Data of treated tibialis anterior muscle was compared with untreated tibialis anterior of each individual (Figure 4). A two-fold increase in gene expression was considered upregulated expression. Gene expression of IL-6 and TNFα was in none of the three participants >2-fold increased due to MABs delivery. Only expression of chemokine SDF1a was >2-fold increased in the treated muscles of participant A and B. In addition, analysis of H&E stained muscle samples by a pathologist verified that MABs administration did not result in an increased presence of leukocytes in the MABs-treated leg. In summary, autologous MABs administration does not result in a skeletal muscle inflammation response.

**Figure 4.**
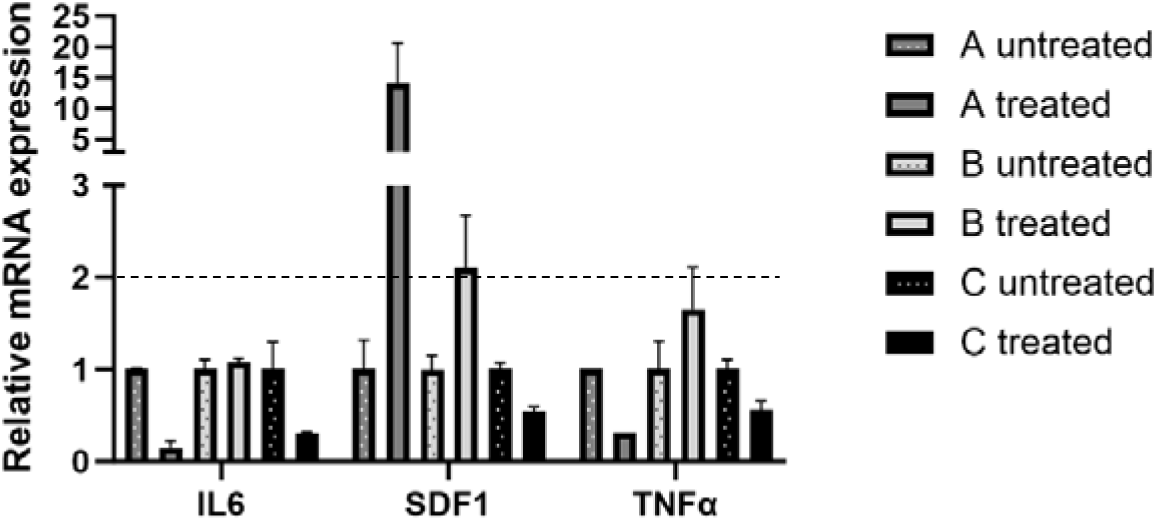
Assessment of skeletal muscle inflammation following ATMP delivery Relative mRNA expression in treated and untreated tibialis anterior muscle biopsy at visit 5 of SDF1a (a), TNFa (b) and IL-6 (c) was normalized to TBP housekeeping gene. Expression of treated sample was compared with individual’s untreated sample, a >2-fold change is considered increased. One muscle sample was collected from left and right TA muscle from each participant for RNA isolation and cDNA isolation, qPCR was performed in triplicate.

### Secondary objectives: skeletal muscle homing and muscle regeneration

The secondary objective of the phase I/II clinical trial was to assess MABs homing and induction of muscle regeneration in tibialis anterior muscle of the treated leg. To stimulate homing, 1 minute and 40 seconds bouts of dorsiflexion exercise were performed with both legs till exhaustion on a Biodex device. As shown in supplementary figure S1, a 36-42% reduction in the cumulative amount of physical work performed (total work) of left leg was observed in the last set compared to first set of subject A and C. Subject B did not show a difference in total work (Joules (J)) between first and last exercise bout, even though she performed more bouts than subject A and C, of which a number showed >40% reduced total work (J). Assessment of migration of ICG-labeled MABs was not possible, as ICG labeling was discontinued after adverse event in participant B and no muscle biopsies were taken due to the discomfort. Alternatively, we assessed MABs migration by quantification of the total number of NG2+ cells, as marker for both residing and administered MABs, per muscle fiber in biopsies of the treated and untreated m. tibialis anterior 24 hours after ATMP administration in participant A and C. As shown in figure 5, a significant increase of 20±7% (p<0.05) in NG2+ cells/muscle fiber was observed in a biopsy of the treated muscle versus the untreated muscle in participant A. In participant C, no significant increase was observed, but fields with increased number of NG2+cells/fiber were observed in muscle of MABs-treated leg. Microscopic analysis of the muscle biopsies collected of both treated and untreated m. tibialis anterior 4-6 weeks after MABs administration of all three subjects showed that the mean number of NCAM-positive muscle fibers was increased in all three subjects, but large variation exists between regions in the muscle biopsies (figure 6).

**Figure 5.**
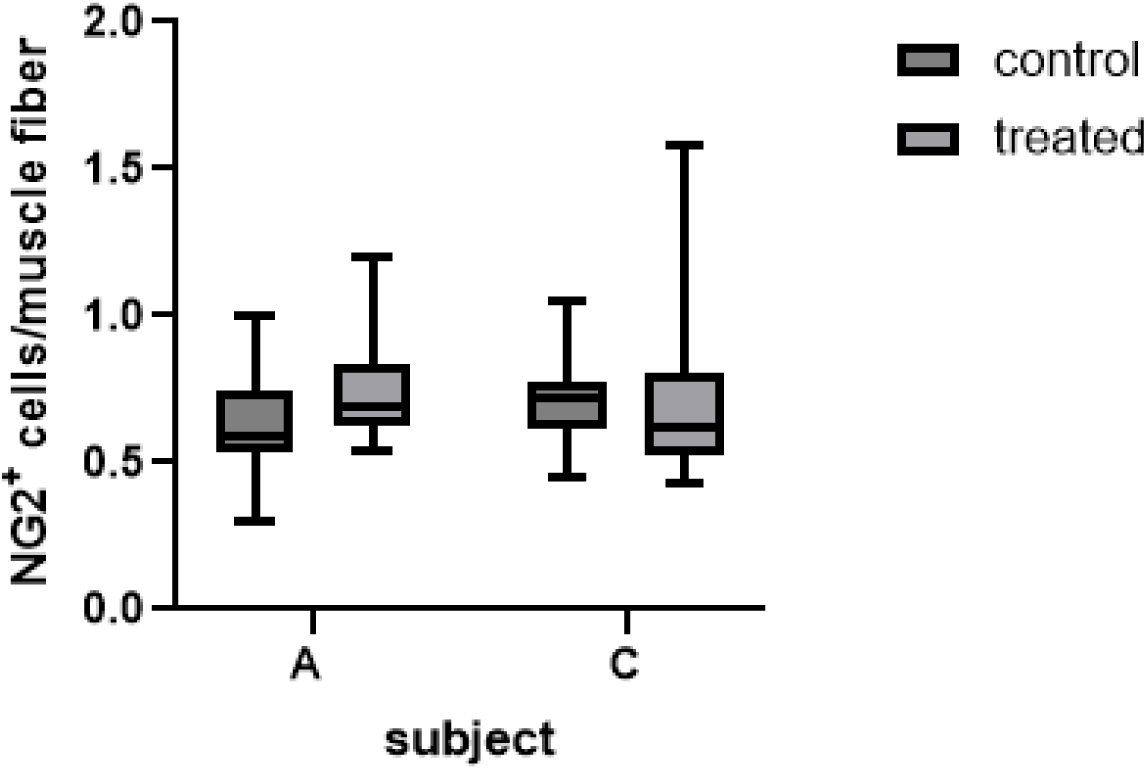
Quantification of NG2+ cells/muscle fiber 24h after i.a. MABs administration in left lower leg in phase I study, a tibialis anterior muscle biopsy of the left (treated) and right (untreated) leg was collected from 2 out of the 3 patients (a and c). Per biopsy, a minimum of 400 muscle fibers were imaged for WGA to quantify the number of muscle fibers/field and NG2+ to count the number of MABs. Counting was performed blinded.

**Figure 6.**
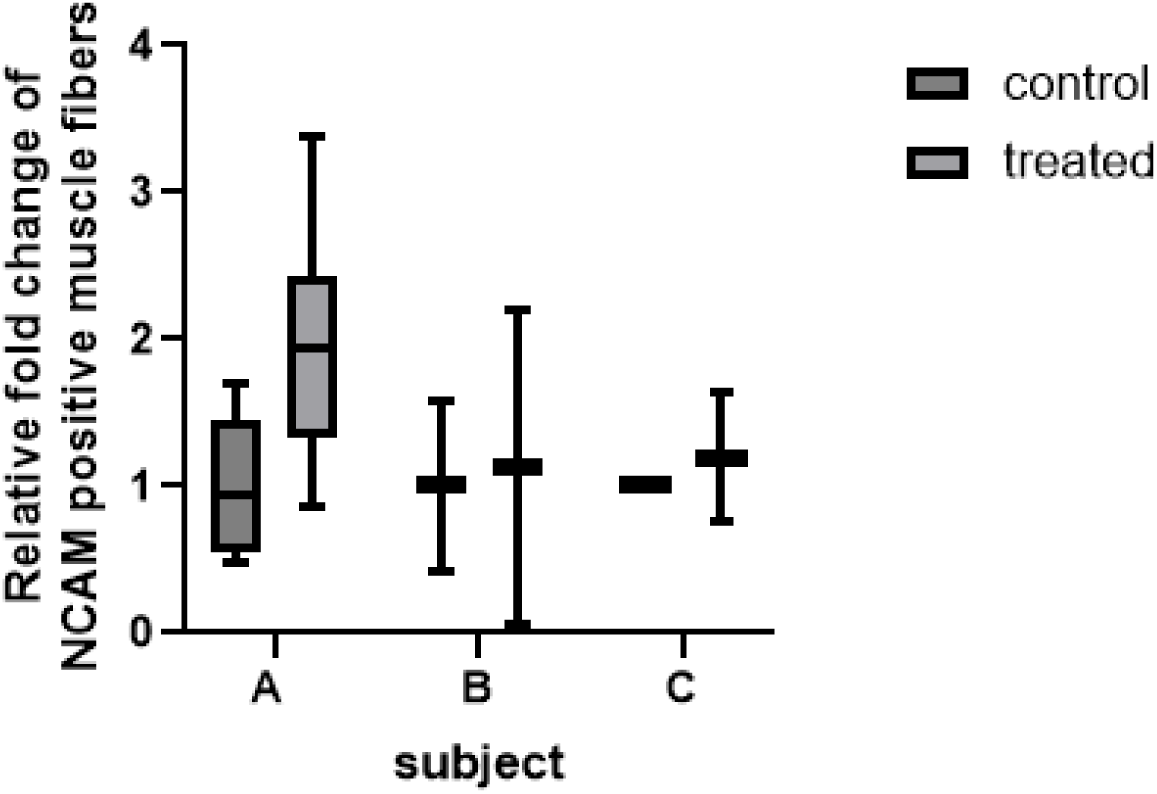
NCAM staining of tibialis anterior (TA) muscle biopsy of treated and untreated leg. Following DAB-NCAM staining, quantification of NCAM positive and negative muscle fibers. Per subject, two muscle biopsies of left and right TA muscle were analyzed, only for subject C only one muscle sample of the right TA (control) was available for quantification. A minimum of 300 muscle fibers were quantified per sample by a blinded researcher. TA: tibialis anterior muscle.

## Discussion

Here we report the results of the first Phase I/II clinical trial of human intra-arterial administration of autologous mesoangioblasts in the tibialis anterior muscle of m.3243A>G mutation carriers to assess primarily safety and secondary efficacy with respect to homing and induction of muscle regeneration. We found that autologous MABs transplantation in tibial artery is safe and well tolerated. Comparison of the muscle biopsy of the treated versus the untreated contralateral tibialis anterior muscle suggests homing and regeneration of administered MABs.

### ATMP production

A total of 50 million MABs/kg body weight of transplanted limb (lower leg 5% of body weight) were transplanted as used by Cossu et al. with allogeneic MABs [13]. For intra-arterial transplantation of one tibialis anterior muscle, sufficient autologous MABs could be cultured in the GMP facility from all three m.3243A>G mutation carriers within 6-7 passages, demonstrating the capability to culture 3.8 billion MABs for future transplantation of all muscle groups in the body within 15 passages, herewith preserving MABs capacity to proliferate *in vivo* following transplantation [12, 19]. As autologous MABs were used, the m.3243A>G mutation load was determined in the intermediate product (IP) after MACS sorting at passage 3 and the medicinal product (MP) at passage 6/7. During MABs expansion, the m.3243A>G mutation load increased on average 7-8%, and in one case 15%. This does not lead to safety issues as the likelihood of being unaffected at 27% is around 90% [20], and the mutation load is still >25% below the skeletal muscle mutation load of the individual. This increase in mutation load was not observed when MABs from these participants, collected at another study visit, were cultured under the same conditions in a research lab, nor was this observed in MABs from four additional m.3243A>G mutation carriers (see supplementary figure S3). Changes in the mtDNA mutation load have been reported to occur during culture of primary cells or mtDNA mutation carrier [21, 22], which may be random and cannot be prevented. Based on this limited data, it is not possible to draw conclusions on whether or not this is a consistent phenomenon in a GMP culture or a coincidence. In addition, the variation between replicates and methods for m.3243A>G mutation load analysis needs to be reduced.

### Safety

Our primary aim was to show safety of intra-arterially injecting ∼250 million autologous MABs in the tibial artery. No SAE, no vascular obstructions measured by angiography, and no changes in heart rate, blood pressure, oxygen saturation or breathing frequency were observed during 24h following MABs administration. One grade II local adverse event was observed and resulted in elevated pain score, namely local inflammation in treated lower leg of participant B treated with 10% ICG-labeled MABs in Hypothermosol to enable quantification of autologous ICG-positive MABs in the tibialis anterior muscle biopsy. The excipient solution Hypothermosol is FDA approved and has been used in many clinical studies using various administration routes. ICG binds to plasma proteins and is amongst others used for measuring cardiac output and to visualize blood vessels and lymph nodes during surgeries. Reported adverse reactions of ICG include urticarial reactions [23], which were observed in participant B. Since it is not possible to discriminate if the adverse event is caused by the ICG or by sensitivity of participant B to (other components of) the MABs administration, we decided to discontinue the ICG and participant A and C were treated with unlabeled MABs. In contrast to the inflammation of treated left lower leg of patient B, the front side of the treated m. tibialis anterior felt a bit warm in participant A and C, and a temporal increase in plasma IL-6 and TNFa at 8h and 16h post administration of all participants may be responsible for this slight elevation of 1,17±0,15°C in body temperature. The direct and short window of increased IL-6 is likely linked to cellular stress due to the ATMP delivery procedure. In contrast, SDF1a was not increased in plasma following intra-arterial transplantation, but increased in skeletal muscle biopsy of treated tibialis anterior muscle 4-6 weeks after MABs administration in participants A and B. SDF1a expression is associated with increased muscle regeneration [24], but it would require further investigation to show that this is the cause in these participants.

No immune response was observed in the current study. This was expected as autologous MABs were transplanted and since wild-type tRNAleu is present in all patients with the heteroplasmic m.3243A>G mutation. This suggests that mtDNA mutation carriers are more suitable candidates for cell therapy compared to carriers of a nuclear DNA mutation(s) that completely lack the wild-type protein. For example, antibodies against wild-type Dystrophin protein have been observed upon transplantation in patients with DMD [25]. Taken together, intra-arterial administration of autologous MABs as ATMP is safe, but all patients experienced some discomfort, which manifested from very mild (patient A) to more severe (patient B), which could be an individual-specific reaction to the ATMP and procedure or related to IC-Green. Since this is only the second clinical study where MABs are administered intra-arterially, and the first study using autologous MABs, further research is needed to determine the cause of the discomfort. In a previous DMD trial, donor MABs were administered under general anesthesia [12], making it unclear whether the discomfort observed is specific to autologous MAB administration, MAB administration in general, or related to the ATMP administration procedure itself. For example, it is worth investigating if adjusting the procedure, such as administering the ATMP at a slower rate or pre-warming it to 37°C, might reduce the discomfort.

### Homing and regeneration capacity

The secondary objective was to assess migration and regenerative capacity of autologous MABs upon intra-arterial delivery. Muscle damage and the resulting inflammation is required for MABs migration from the artery to the muscle [10] and can be present due to the disease, e.g. in muscular dystrophies, or can be triggered by inducing muscle damage, e.g. cardiotoxin injection in mice [26]. In humans, eccentric exercise induces muscle damage, which can be measured by the expression of infiltration markers. Also, Paulsen et al. demonstrated inflammation and leukocyte infiltration in exercised muscles following exercise-induced 40-50% reduction of maximum strength [16]. As muscle damage is generally not present in m.3243A>G carriers, including the subjects in this study, dorsiflexion exercise sessions till exhaustion were performed with both lower legs 24hrs before i.a. MABs administration. Although, we did not quantify peak torque before and after the eccentric exercise, on average, total work (J) was reduced by 26±8% (mean±SEM) in the last session of 1 minute 40 seconds compared to the first session it is reasonable to expect that muscle damage and local inflammation was induced in our study. To assess MABs homing, quantification of NG2+ cells in muscle biopsies collected 24h after administration, which has been described in literature for MABs quantification [27, 28]. Quantification of NG2+ cells per muscle fiber showed a significantly higher number of MABs in the transplanted TA muscle of individual A, while individual C showed some fields with increased NG2+ cells, but this was not significantly different from the untreated TA muscle. This data suggest homing of i.a. transplanted autologous MABs, but without labeling we cannot discriminate between autologous transplanted and residing MABs. Torrente et al. quantified intra-muscularly injected autologous CD133+ cells in muscle cryosections using Prussian blue staining of iron dyne beads that were incorporated upon CD133+ MACS sorting [25]. This approach was successful for assessing intra-muscularly injected cells, but requires availability of a GMP-grade antibody coated magnetic beads. In addition, a biopsy may not represent the entire muscle upon intra-arterial administration of stem cells. To evaluate the homing of systemically administered stem cells and assess their distribution overall muscle groups, it is essential to develop GMP-compliant cell labeling procedures that provide sufficient signal intensity for non-invasive detection methods.

## Conclusion

Taken together, this phase I/II clinical study demonstrated that intra-arterial delivery of autologous MABs as ATMP is safe and well tolerated. Data with respect to homing of the mesoangioblasts to the transplanted muscle were promising, but not conclusive. Based on animal studies, minimal three mesoangioblast doses at 4–6-week interval are needed to achieve functional improvement. This will be the aim of the next phase IIa intra-subject controlled study (NCT05962333).

## Declarations

### Ethics approval and consent to participate

This intra-subject controlled mono-center study was approved by the Central Committee on Research Involving Human Subjects (CCMO) in the Netherlands (NL68732.000.19, June 7, 2020), was conducted according to the principles of the Declaration of Helsinki and in accordance with the Medical Research Involving Human Subjects Act (WMO) and other guidelines, regulations and acts.

### Consent for publication

All participants in this study have provided written informed consent for the publication of their anonymized data and related information.

### Availability of data and materials

The data that supports the findings of this study are available from the corresponding author upon reasonable request.

### Authors’ contributions

FT: Study conception and design, execution and data collection, analysis and interpretation of results, manuscript preparation and funding acquisition. JH and CL: Execution and data collection, manuscript revision. CL and ET: Data collection, analysis and interpretation of results and manuscript revision. NW: Execution and data collection, manuscript revision. PL: Analysis and interpretation of results, manuscript revision. FY: Analysis and interpretation of results. IW, SK, PM, FL: Execution and data collection, manuscript revision. HR and LW: Execution and data collection. IC: Study conception and design, execution and data collection, analysis and interpretation of results, manuscript revision. HS: Study conception and design, analysis and interpretation of results, manuscript revision and funding acquisition.

## Acknowledgements

The authors would like to express their outmost gratitude to the m.3243A>G mutation carriers that participated in this study, without whom this study would not have been possible. We would also like to thank all monitors for observing the participants for 24h after transplantation, and support staff from pharmacy, medium care and intervention radiology Maastricht UMC, and all other persons who contributed to this study.

