## Supplemental material for "Intra-arterial transplantation of autologous mesoangioblasts in m.3243A>G mutation carriers is safe – first phase I/II human clinical study"

**In- and exclusion criteria**

*In order to be eligible to participate in this study, a subject must meet all of the following criteria:*

-	Written informed consent

-	Age: 18+

-	Sex: male/female

-	Patients with the m.3243A>G mutation

*A potential subject who meets any of the following criteria will be excluded from participation in this study:*

-	Use of anti-coagulants, anti-thrombotics and other medication influencing coagulation

-	Have a weekly alcohol intake of ≥ 35 units (men) or ≥ 24 units (women)

-	Current history of drug abuse

-	Deficient immune system or autoimmune disease

-	Significant concurrent illness

-	Ongoing participation in other clinical trials

-	Major surgery within 4 weeks of the visit

-	Vaccination within 4 weeks of the visit

-	Pregnant or lactating women

-	Psychiatric or other disorders likely to impact on informed consent

-	Patients unable and/or unwilling to comply with treatment and study instructions

-	Any other factor that in the opinion of the investigator excludes the patient from the study

-	A history of strokes

-	Allergy for contrast fluid

-	Peripheral signs of ischemia or vasculopathy

**Table S1**. Conditional release criteria of 3 MP batches of autologous MABs

| Description | | Method | Release Criteria | Subject a  (MAB001 lijn 2) | Subject b  (MAB002) | Subject c  (MAB003) |
| --- | --- | --- | --- | --- | --- | --- |
| Cell number | | Trypan blue hemacytometer | 5*10E6/ml * 40 ml^**^ | 175*10E6 in 35 ml (5*10E6/ml) | 145*10E6 in 29 ml (5*10E6/ml) | 170*10E6 in 34 ml (5*10E6/ml) |
| Cell viability | | Trypan blue hemacytometer | >70% | 88,8% | 97,4% | 90,4% |
| Definitive result sterility start material | | Bactec | Negative | Negative | Negative | Negative |
| Definitive result sterility intermediate product | | Bactec | Negative | Negative | Negative | Negative |
| Definitive result sterility final product (MP) | | Bactec | Preliminary result Negative | Preliminary result Negative | Preliminary result Negative | Preliminary result Negative |
| Immuno-phenotype | **CD34**  **CD31**  **CD44**  **CD13**  **CD56**  **CD45** | FACS | ≤ 5%  ≤ 5%  ≥ 90%  ≥ 90%  ≤ 5%  ≤ 5% | 0,11%  0,2%  99,8%  99,4%  0,07%  0,12% | 0,14%  1,81%  99,9%  98,1%  0,54%  0,70% | 0,14%  0,72%  99,8%  99,3%  0,15%  0,18% |

**Table S2.** Final release criteria of 3 MP batches of autologous MABs

| Description | Method | Release Criteria | Subject a | Subject b | Subject c |
| --- | --- | --- | --- | --- | --- |
| Mycoplasma test | PCR | Negative | Negative | Negative | Negative |
| Microbiological control of cellular product | Bactec | Negative | Negative | Negative | Negative |
| Endotoxin | LAL assay | ≤1 EU/ml | ≤1 EU/ml | ≤1 EU/ml | ≤1 EU/ml |
| Patient verification | Genescan analysis | 100% match | 100% match | 100% match | 100% match |
| Mean mtDNA mutation load | Genescan analysis | <15% | 18%* | 27%* | 23%* |

* Out of specification;

**Table S3.** Primers qPCR

|  | Forward (5’-3’) | Reverse (5’-3’) |
| --- | --- | --- |
| TBP | CACGAACCACGGCACTGATT | TTTTCTTGCTGCCAGTCTGGAC |
| IL-6 | GGTACATCCTCGACGGCATCT | GTGCCTCTTTGCTGCTTTCAC |
| SDF1a | GTGGTCGTGCTGGTCCTC | AGATGCTTGACGTTGGCTCT |
| TNFa | GACAAGCCTGTAGCCCATGT | GAGGTACAGGCCCTCTGATG |

**Table S4.** m.3243A>G mutation load in MABs GMP culture phase I/II clinical study

|  | IP Pacbio | MP Pacbio | MP genescan |
| --- | --- | --- | --- |
| a | 6% | 18% | 11±1% |
| b | 12% | 27% | 32,5±3,5% |
| c | 16% 1^st^ run  18% 2^nd^ run | 23% | 26,5±3,5% |

IP: intermediate product after MACS sorting; MP: Medicinal Product; MP2: 2nd MP sample; PacBio m.3243A>G mutation load analysis is NGS-based analysis performed by Clinical Genetics department as IP and MP release test. Genescan analysis of m.3243A>G mutation load was performed in duplicate following diagnostic SOP for m.3243A>G mutation load analysis, but performed in a research lab.

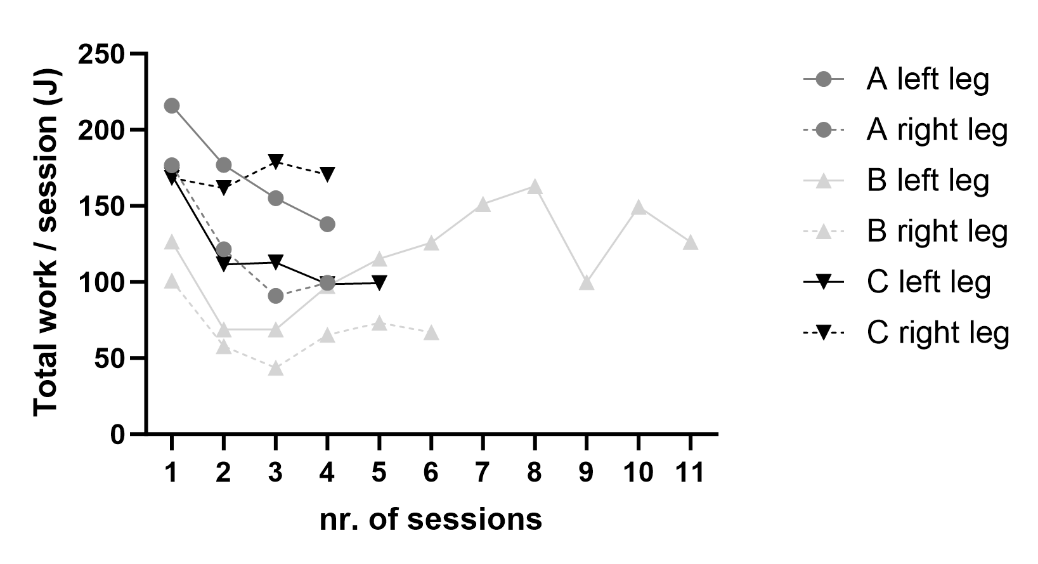

**Figure S1. Eccentric exercise of lower legs 1 day before i.a. delivery**

A maximum bout of eccentric exercise on a Biodex was performed with both lower legs 1 day before intra-arterial delivery of autologous MABs. Sessions of 1 minute 40 seconds were performed, followed by a 30 second break until exhaustion. Exercise was first performed with the left lower leg and the same number of sequential sessions was performed with the right leg, if possible.

| a | 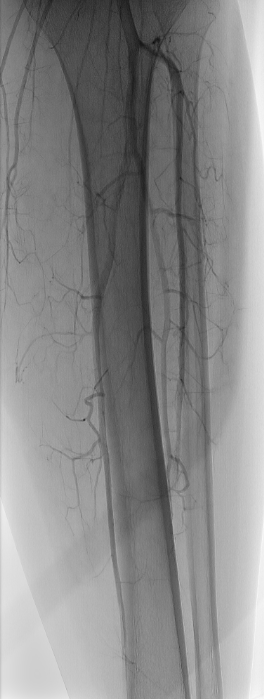 | b | 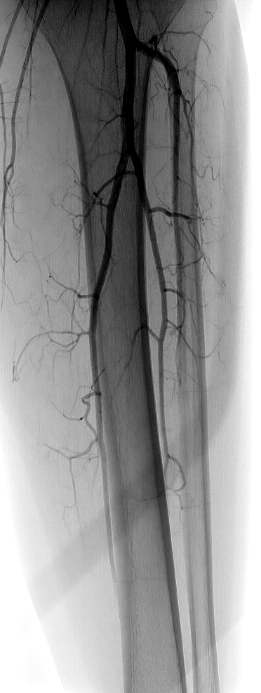 | c | 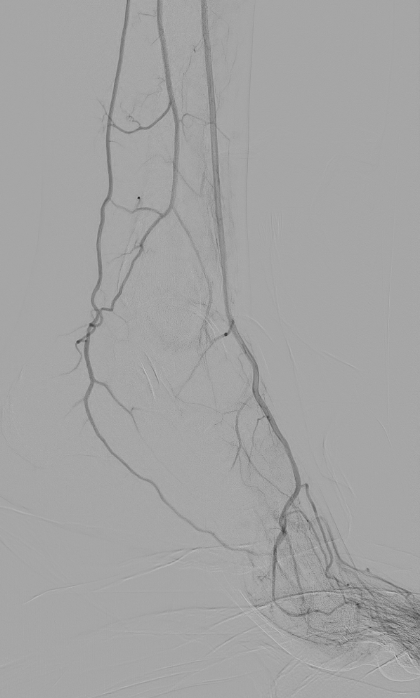 |
| --- | --- | --- | --- | --- | --- |
| **Figure S2. Angiographic evaluation pre and post MABs infusion.**  a) Angiography of left lower leg before MABs infusion, showing patency of popliteal and tibial arteries; b) Angiography of left lower leg after MABs infusion, confirming patency of popliteal and tibial arteries; c) Angiography of left foot after injection confirms patency of dorsalis pedis and plantar arteries. | | | | | |

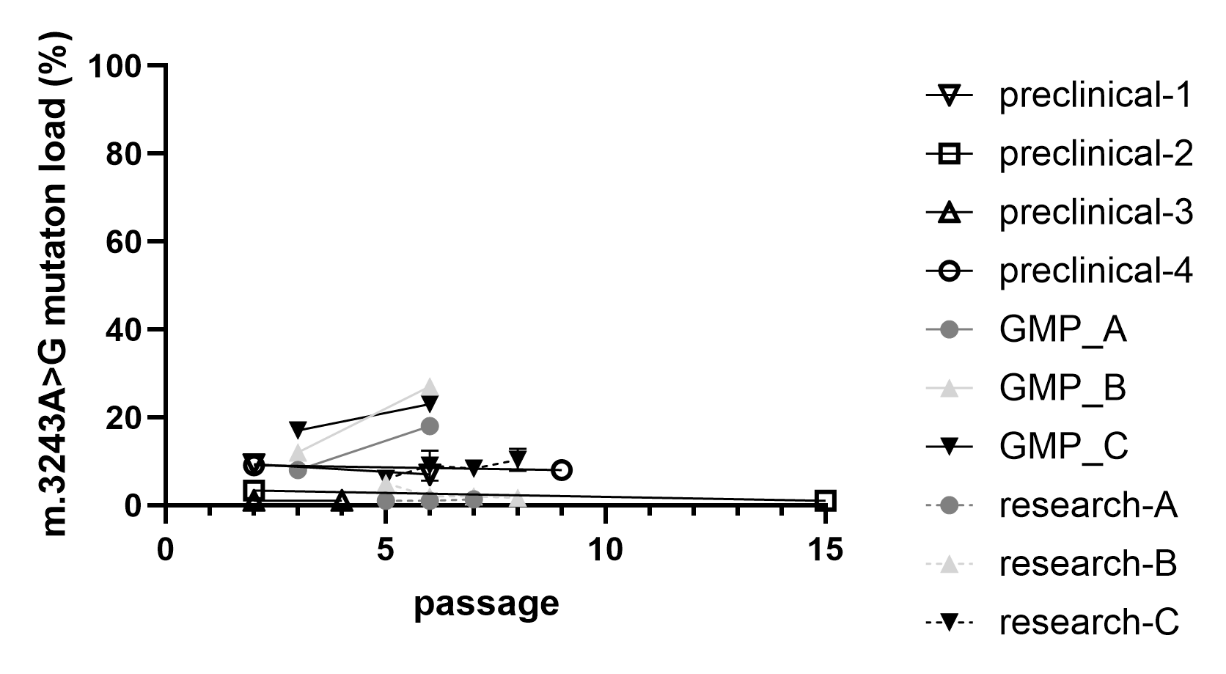

**Figure S3. m.3243A>G load analysis at different passages during MABs culture.**

Quantification of m.3243A>G mutation load of mesoangioblasts of participant A, B and C collected at visit 1 that were cultured in the research lab (research- ), mesoangioblasts of participant A, B and C collected at visit 2 that were cultured in the GMP lab and transplanted (GMP_ ), and four additional mesoangioblast cultures that were analyzed in the research lab during preclinical phase (preclinical-).
